# PREVALENCE OF HCV AMONG HOUSEHOLD MEMBERS OF HEPATITIS C VIRUS-INFECTED PATIENTS ATTENDING CENTRE UNIVERSITAIRE DE SANTE PUBLIQUE (CUSP) HEALTH CENTER

**DOI:** 10.1101/2024.12.27.24319696

**Authors:** Patrick Nemeyimana, Uwumuremyi Fabrice, Ruhumuriza anselme, Bakunzibake Pierre, Emmy Mugisha, Valens Karenzi, Eliah Shema, Pascal Kiiza

## Abstract

Globally, hepatitis C is included in the public health concern. More than 1.4 million deaths are reported. In 170 million who are infected with HCV worldwide,71 million have serious complications. There is a close relationship between hepatitis C infections among family members with one case. Family member’s behaviors and exposure to positive family members without precautions are among the risk factor of getting hepatitis C infection. In Rwanda, the burden of Hepatitis C infections reaches to 4% of infected people. High prevalence of hepatitis C infection can be associated with horizontal transmission among household of positive patients for HCV. In Rwanda no data showing the contribution of horizontal transmission of HCV among household members as one of the risk factors to increase the prevalence of HCV.

The objective of this study was to determine the prevalence of HCV among the household members living with HCV positive individual, specifically attending centre universitaire de santé public of BUTARE.

The study used a descriptive cross-sectional design to assess the prevalence among 30 families having had positive cases for HCV with 180 sample size. With use of questionnires, HCV risk factors were assessed from our participants and from their responses. Purposive sampling was done for HCV rapid test and positive cases sample were confirmed at university teaching hospital of BUTARE with Elisa.

From participants tested, 10.8% were tested positive for HCV counting for the prevalence of our study. Our findings are in line with other same studies conducted in Egypt and Iran revealed that there was 14.1%, which is a bit closer to our finding. we have found that unprotected sex with parnters and marital status between infected cases has a big influence to the high prevelence.

Based on the findings from our study, 10.8% of our participants tested positive for HCV due to limited education, awareness, and exposure to contributing risk factors about HCV transmission.

## CHAPTER 1: INTRODUCTION

### 1.1. BACKGROUND

Hepatitis C viral infection is among the major global public health problems. Annually about 1.4 million die from this infection due to its complications including cirrhosis and liver cancers.(Jefferies *et al*., 2018). Globally, people who have HCV infection are more than 170 million and of whom, 71 million have chronic hepatitis C virus infection (Dhawan *et al*, 2019)

In Rwanda, About 4 % have been were found to be infected by that virus(James and Nkurunziza, 2019). In study conducted in Egypt, it has revealed that 35.6% had infected with HCV due to living infected persons which is higher than 5.2% who had infected without contact from known positive cases in their families(Omar *et al*., 2017). The appearance of this viral diseases lead to severe complications including liver cirrhosis, hepatocellular calcinoma,liver fibrosis and when it is not managed can lead to morbidity and mortality rise (Case-lo, 2020). Role of horizontal household transmission was also previously reported by other authors from Egypt and other countries like Iran and showed an increased prevalence.

There is a close relationship between hepatitis C infections among family members with one case. Family member’s behaviors and exposure to positive family members without precautions are added risk factor of getting hepatitis C infection(Indolfi, Nesi and Resti, 2013). This has been supported by the study conducted in Egypt that horizontal intra-familial transmission plays a big role in increasing the prevalence of hepatitis C infection (Mz *et al*., 2020).

Exposures including saliva, blood contacts and sharing of needles were among the risk factors to acquiring this infection from family members, in addition to parenteral children birth. This was found especially among the family members of chronic hepatitis patients(Wiley and Strickland, 2020).Both sexual and nonsexual practices can be risk factors for hepatitis C infection. Among the household members, not only the sexual but also sharing of toothpaste and razors can increase the risk(Hajiani *et al*., 2020).

Recommended by this author is that assessing horizontal intra-familial transmission of hepatitis C infection would assist in its prevention. This support the target of Minister of health to eliminate hepatitis C infection by2030 (Musabeyezu., 2020).

HCV infection to be eliminated, enough screening and treatments need to be afforded by patients. Our study of assessing the prevalence of HCV among household members living with positive patients will facilitate the ministry of health strategies set to eliminate this infection among community.

## OBJECTIVES

### GENERAL OBJECTIVE

To evaluate whether horizontal transmission of HCV among household can contribute to the increase of HCV prevalence.

### SPECIFIC OBJECTIVES

1. To determine the contribution of horizontal transmission of HCV among household can increase prevalence
2. To compare and rate the risk factors of HCV transmission among household members.
3. To assess the awareness of HCV transmission among participants.

### RESEARCH QUESTIONS

**1.** What is the contribution of horizontal household HCV transmission in HCV positive patients attending CUSP to the increase of HCV prevalence?
**2.** Which among risk factors mostly contribute to the household transmission of HCV?
**3.** How is the awareness of people about transmission and prevention ways of HCV?

### SIGNIFICANCE OF THE STUDY

This study will provide the data for the household transmission of HCV which will help in its management and being precautioned about its transmission ways which will help to minimize its prevalence.

To contribute to the elimination program of HCV in Rwanda by giving statistical data from our study.

To participants who will be tested positive their records and statuses will be submitted to the CUSP health center for further fellow up. This will support in HCV elimination target by Rwandan ministry of health

### MOTIVATION OF STUDY

This study is the significant willing to participate in the elimination of hepatitis C infection in Rwanda adding a step to the goal of the ministry of health and to transmission among the patients.

Studies from other countries have shown that family history plays a key role in the transmission of this infection that pushed us to concentrate more on the household members living with positive patient. So that the data can be made available on the correct number of people who are HCV positive.

The information that our study will provide will be based on to set strategies so that family members living with such patients will be taken into account for early screening of household members of a positive case for HCV in order to reduce transmission and limit the complications leading to eliminating this infection.

## CHAPTER 3. METHODOLOGY

### 3.1. INTRODUCTION

This chapter describes the methodology we used for the study; Study area, study design, study population, sample size, sampling strategy, data collection methods, data analysis, limitation of the study and ethical consideration.

### 3.2. STUDY AREA

The study was carried out at CUSP located in the southern province, HUYE district and Ngoma sector in Butare city.

### 3.3 STUDY DESIGN

A descriptive cross-sectional design was used to describe the evaluation of household transmission of hepatitis C among households of HCV positive patient attending CUSP.

### 3.4. STUDY POPULATION

The study was conducted on households’ members of HCV positive patients attending CUSP

#### 3.4.1 INCLUSION CRITERIA

All susceptible household members including spouses, breastfeeding to a positive mother and very close individuals of HCV positive patients ‘attending CUSP.

#### 3.4.2 EXCLUSION CRITERIA

- House hold members who don’t live with any HCV positive index patients.
- Short time family visitors.

### 3.5. SAMPLE SIZE

The study sample size was 180 participants who met the acceptance criteria.

The calculation of the sample size was determined using formula(Rosner, 2010):

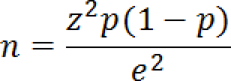

Where

n=minimum sample size

z=standardized normal deviation which is equally to 1.96 (at 95% confidence interval)

P= estimated population prevalence equal to 13.6% (El-bendary *et al*., 2014) ( this prevalence used in calculation is a specific for household members living with positive cases, while 4% as stated in background of study was the general prevalence of HCV in Rwanda, for no national data available for our study, on local data concerning our study was considered.

e= estimated error equal to 0.05

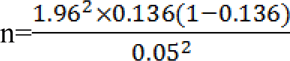

n=180

### 3.6. SAMPLING STRATEGY

Patients were first tested with HCV ab rapid test. ELISA was used as a confirmation test for the ones who tested positive.

### 3.7. DATA COLLECTION METHOD AND PROCEDURE

This study was conducted at Centre Universitaire de Santé Public health centre. The HCV Ab rapid test positive samples were transported in portable cool box and analyzed in immunoserology unit at CHUB hospital for ELISA confirmation. Consent form was designed and questionnaire with multiply choice questions for the participants in order to assess the risk factors and awareness. The form had space to record the results from rapid test and detection from ELISA.

Samples were taken by finger sticking and the test insert form was used for HCV Ab rapid testing. For reactive cases on HCV Ab rapid test, were collected by venous puncture in dry test tubes for confirmation with enzyme linked immunosorbent assay (Elisa). Their results were informed to the owners immediately and confidentially. This study had taken a period of four weeks from 2^nd^ January 2021 to 2^nd^ February 2021 for data collection and analysis.

#### Materials

Personal protective equipment’s (Gloves, Lab coat), testing materials (HCV Ab rapid test kits), blood sampling materials (Needle, dry Test tube, Cotton(pad), Vacutainer, Marker, Phlebotomy chair, Logbook, a tube holder). Waste containers (Sharp container waste bin, infectious Waste bin) and a portable cool box for sample transport.

##### 3.7.1. SPECIMEN COLLECTION

After getting permission from the Head of CUSP to use records on the infected HCV patients, we contacted health care workers from our patients’ cell. They helped us in the mobilization process and the contacted participants met us at CUSP health center. For older ones who were not able to attend, we managed to go to their places to collect their samples. Before collecting samples, we first explained to all participants about our research, its purpose, and the role that it will play to the society. We also explained to them their role and contribution to the success of our study. We proceeded with a brief explanation of how the samples are collected and possible side effects. For those who accepted, they signed the consent forms and were tested on HCV Ab rapid test. We gave them their results, and for the ones tested positive we took other samples for ELISA confirmation with the help of a thermos for their transportation.

##### 3.7.2. TESTING PROCEDURES AND THEIR PRINCIPLES FOR SCREENING AND CONFIRMATORY TESTS OF HCV

###### PRINCIPLE

this test contains nitrocellulose membrane which is precoated with recombinant HCV capture antigen at the test line region. The protein A-colloid gold conjugate and the specimen moves along the membrane chromatographically to the test region. There the antigen-antibody protein a gold particle complex forms into a visible line with high degree of sensitivity and specificity. It has ‘T’ and ‘C’ representing testing and control line.

###### PROCEDURE

-Firstly, PPE were worn and working bench were prepared

-All kit components were put on the bench

-The test devices were removed from foil pouch and placed on flat dry place, labelled with patient identifier.

-fingertip was cleaned using alcohol pad, squeezed the fingertip and lanced with lancet and dispose it in safety box.

-10µl of whole blood were placed on specimen pad on kit.

-4 drops of assay diluent were added

-results were interpreted within 5-20min

###### RESULT INTREPRETATION

➢ NON-REACTIVE: were indicated by the presence of only on control line
➢ REACTIVE: were indicated by the presence of control line and testing line, regardless of which line appeared first.
➢ INVALID: were indicated by no visible line (standard diagnostic inc.2020).

**ELISA TEST (CONFIRMATORY TEST)**

**PRINCIPLE:** ELISA (enzyme-linked immunosorbent assay), is a method that detect antigen/antibody capture in samples using specific antigen/antibody, the quantification is done using enzyme substrate to give color, and the color detected is proportional to the concentration in the sample.

It has four different types and we only used one test (sandwich) in our study (Occurrences, Standard and Linearity, 2010)

### 3.8. DATA ANALYSIS

Data were analyzed using descriptive statistical analysis, Microsoft excel and SPSS and results were presented using frequency table with percentages.

### 3.10. ETHICAL CONSIDERATION

The permission to conduct the study was obtained from the Institutional Review Board, UR-CMHS. Then we were given the permission to carry out the study by the ethical committee of CUSP. We gave enough explanations to the laboratory staff members in their respective departments about the study (objectives, duration and importance to participate) and they all cooperated. The household members who agreed on participating signed a consent form and were given a brief explanation on the whole process. The participation was done voluntary and information gathered were kept in confidential system using personal codes in registration logbook and were only communicated to concerned institutions as requested during study permit. These are CUSP health center, Mayor and Director of Health of Huye district.

## CHAPTER 4: RESULTS

The results of our study presented in histogram and tables. 30 families included 148 tested and answered some questions from our questionnaire. The statistical and correlation of HCV prevalence among household members and risk factors were analysed by excel and SPSS. Most of our population were from rural areas of Huye, Mbazi, Ngoma, Tumba sectors and they were between 5-60 years old. The predominant participants were married.

**Figure 1:**
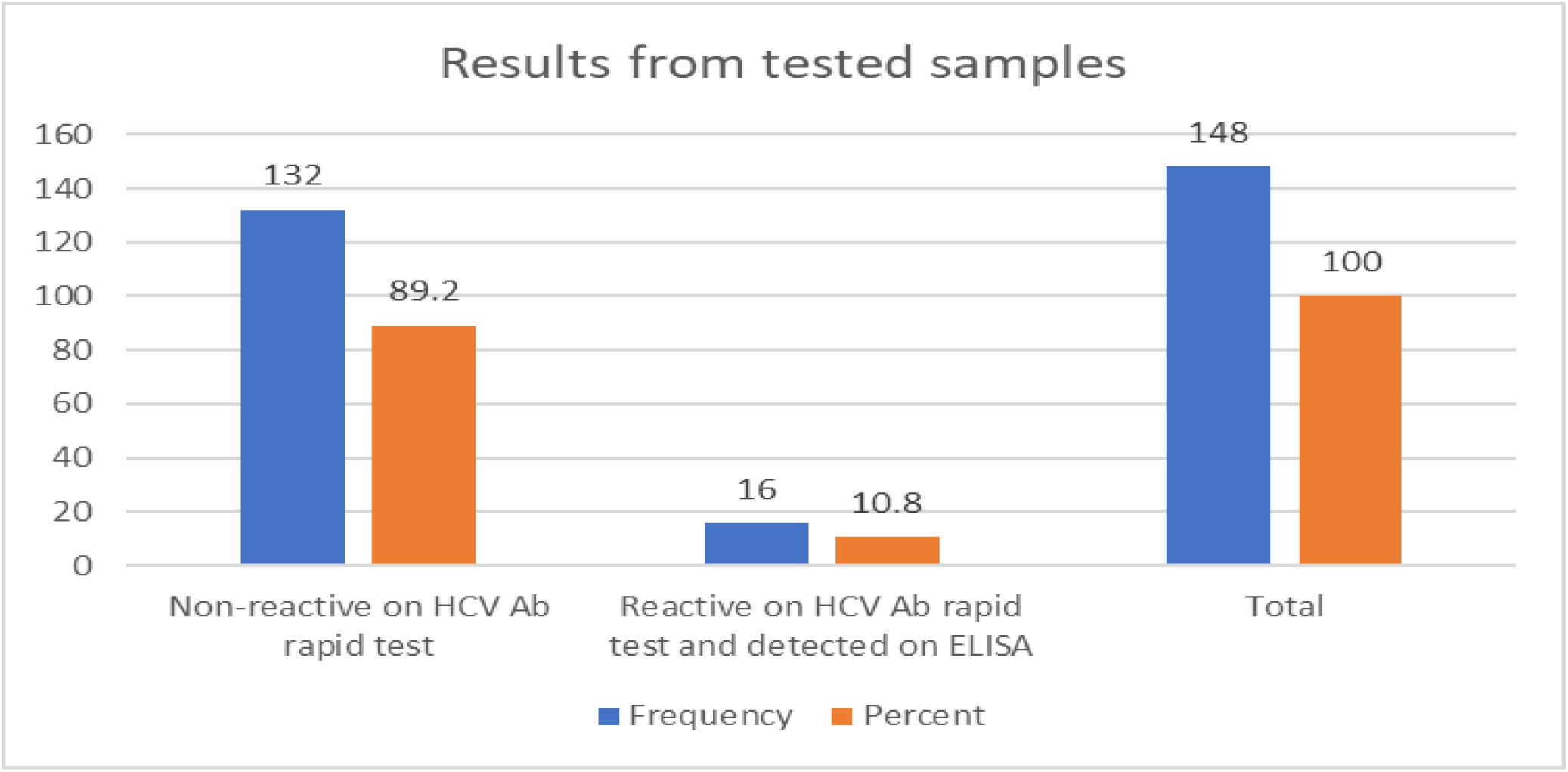
HCV prevalence from the sample collected (blue showing the number of participants and orange showing the rate percentage.

By the use of HCV Ab rapid test and ELISA as confirmatory test. 30 families participated with 148, 16 (10.8%) was reactive on HCV Ab rapid test and detected on ELISA and, 132 (89.2%) were non-reactive on HCV Ab rapid. The prevalence of HCV in our study was 10.8%.

**Table 1:** Association between the HCV risk factors with obtained test results.

| Risk factors | Descriptive | The results from the sample collected |  | Total | p-value |
| --- | --- | --- | --- | --- | --- |
|  |  | Non-reactive on HCV rapid test | Reactive on HCV rapid test |  |  |
| HBV | Yes | 1(0.7%) | 6(4.1%) | 7(4.7%) | 0.001 |
|  | No | 82(55.4%) | 4(2.7%) | 86(58.1%) |  |
|  | Unknown | 49(33.1%) | 6(4.1%) | 55(37.2%) |  |
| HIV | Yes | 0(0.0%) | 4(2.7%) | 4(2.7%) | 0.001 |
|  | No | 132(89.2%) | 12(8.1) | 144(97.3%) |  |
| Blood transfusion | Yes | 2(1.4%) | 2(1.4%) | 4(2.7%) | 0.058 |
|  | No | 130(87.8%) | 14(9.5%) | 144(97.3%) |  |
| Unprotected sex | With my partner | 27(18.2%) | 8(5.4%) | 35(23.6%) | 0.016 |
|  | Other people | 77(52.0%) | 4(2.7%) | 81(54.7%) |  |
|  | No | 28(18.9%) | 4(2.7%) | 32(21.6%) |  |

148 participants tested for HCV, 6 with HBV history tested positive for HCV and 4 who were negative for HBV tested positive for HCV. The p-value =0.001 (The p.Value <0.05 show significant correlation).

4(2.7%) participants who had HIV history tested positive for HCV, this showed high correlation with HCV. with p-Value =0.001.

Among 116 (78.3%) were exposed to unprotected sex, 12(8.1%) people tested positive for HCV and 32 (21.6%) didn’t practice unprotected sex, 4(2.7%) tested positive for HCV and 28(18.9%) were non-reactive on HCV rapid test. The p-Value of this risk factor (P=0.016). This shows a significant correlation with the increase of HCV infection due to unprotected sex.

**Figure 2:**
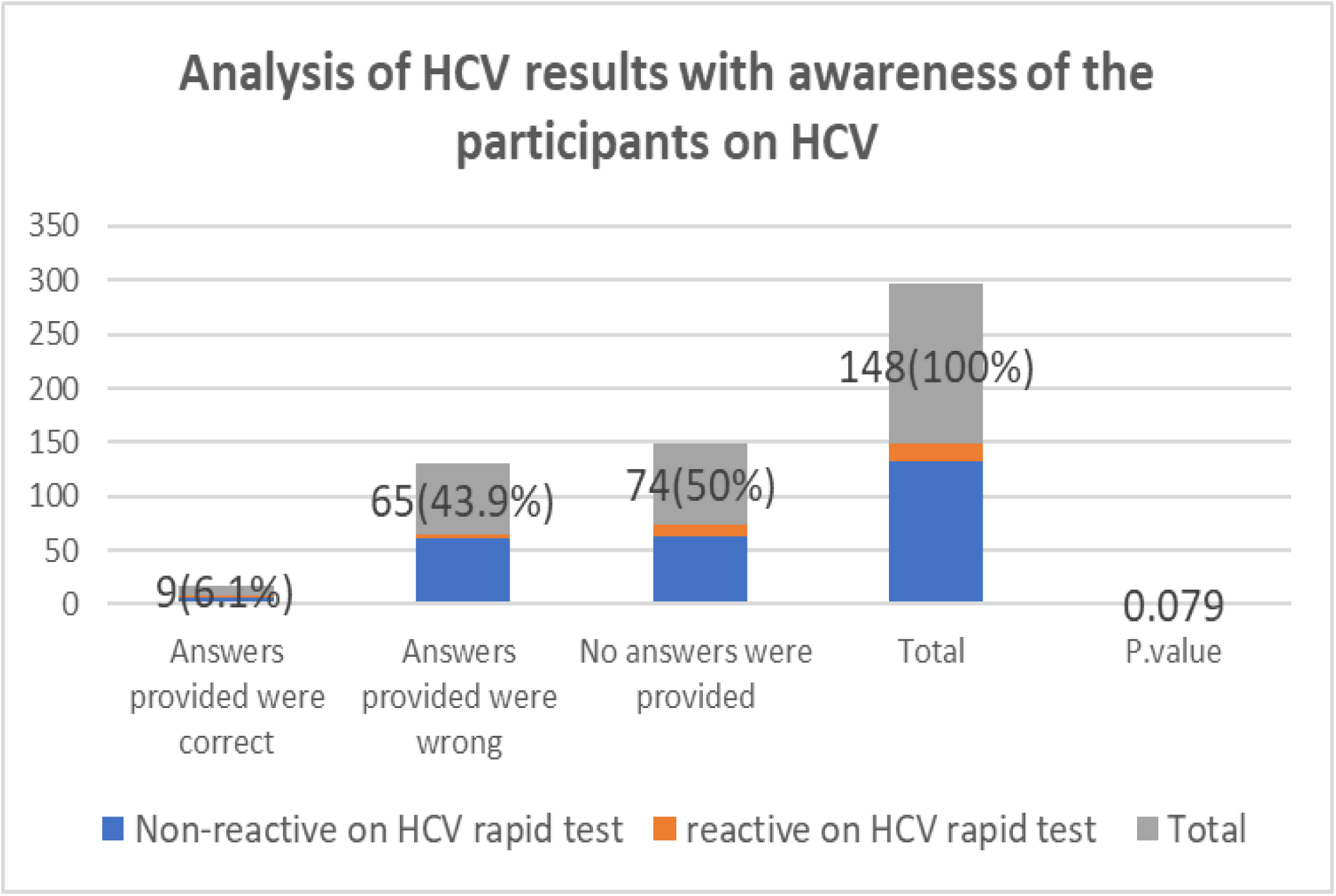
Evaluation of the HCV awareness in comparation with the results. presents the statistical knowledge and awareness of HCV infection transmission. 139(93.9%) participants provided wrong answers, 9(6.1%) provided correct answers. The p-value= 0.079, it is not significant (p-value <0.05 indicates significance). From these results, the awareness about the HCV transmission is very low among our participants.

## CHAPTER 5: DISCUSSION

Hepatitis C is a global burden though it still taken lightly by some people it causes serious complications; liver fibrosis, cirrhosis, hepatocellular carcinoma as result to that complication it leads to morbidity and mortality (Omar *et al*., 2017).

In Rwanda, the reported general prevalence for HCV is 4%. Makuza et al highlighted a strong association between risk factors; old age, unprotected sex, exposure to blood and blood products with this prevalence. Among the members of the household of HCV positive cases, it has shown that the risk for HCV transmission cause the rise of the prevalence(Lankarani *et al*., 2016).

This study showed that the prevalence of HCV was 10.8% among our participants. Comparing our findings with the study conducted in Egypt and Iran with the prevalence of 14.1 %. This implicates the association of HCV transmission among households’ members living with positive cases. The p-value =0.001. Therefore, based on high prevalence of obtained, earlier testing and follow-up to people living with HCV positive cases is required.

Regards to risk factors, 116(78.3%) participants practising unprotected sex either with their partners or other outside partners, 12(8.1%) tested positive for HCV, the p-value=0.016. this has a strong correlation of HCV transmission. This was supported by (Hajiani *et al*., 2020) who stated that ‘unprotected sex is one of a highly contributing risk factor for HCV transmission’. Thus, protected sex is a key in threatening HCV transmission by the use of different ways includes use of condom.

In this study blood transfusion accounted for 1.4% of prevalence with p-value=0.058, no significance (p-value<0.05 shows significance). This risk factor doesn’t show a contribution to the increase of HCV in our study.

About the HCV transmission awareness, the study showed that 125(84.5%) were not aware of HCV and non-reactive on HCV. The P-value for the awareness =0. 079, this shows no significance, (p-value<0.05 shows significance). there is no relationship between the awareness for HCV with the obtained positive result. no correlation between awareness and HCV prevalence as highlighted by (Mz *et al*., 2020),2017. However, basing on population of this study awareness has no contribution in HCV transmission, but people should be mobilized and educated about the transmission of HCV, hence contributing to its prevention.

About HBV and HIV as risk factors for HCV, 6(4.1%) and 4(2.7%) were reactive on HCV respectively. P-value=0.001, it is significant to this study (p-value<0.05 means significance). This was supported by other studies, people with HIV or HBV are immunocompromised hence increase the chance of being infected with HCV (Mbituyumuremyi *et al*., 2018). Therefore, HIV-HCV and HBV-HCV co-infection were highly associated with HCV transmission.

## CHAPTER 6: CONCLUSION, LIMITATION AND RECOMMENDATIONS

### 6.1: CONCLUSION

Based on findings from this study, high prevalence (10.8%) of HCV among household members living with positive case was found. During this study, various risk factors that can contribute to HCV transmission were put into consideration and includes sharing of sharp materials, HIV, HBV, old age, marital status, blood transfusion, unprotected sex as well as limited awareness. Finally, findings from this study revealed that unprotected sex was the most cause of HCV transmission among participants as most of all positive cases were exposed to this factor. Therefore, follow up for HCV positive patient and early HCV screening of family members living with a positive case should be done as a way of preventing intrafamilial transmission.

### 6.2: LIMITATIONS

This study has some limitations. The first one concerns with study style, cross-sectional because we couldn’t know exactly if the positive patient originally got the infection from their household members. Therefore, the phylogenetic study is recommend to confirm the same genotype and subtype in the family to determine the origin of infection.

### 6.3. RECOMMENDATIONS

#### TO THE PARTICIPANTS

Due to high prevalence of HCV among household members, we recommend that family members, with a case of HCV, infected individual(s), they should be educated on how HCV is transmitted, its risk factors and taking initiative for early screening for HCV.

#### TO THE MINISTRY OF HEALTH

our study identified that there is inadequate HCV positive patients follow up which may lead to chronic hepatitis Complications and horizontal HCV transmission. We recommend to the department in charge that upon HCV detection, there should be a follow-up and monitoring to ensure the early starting of treatment and educational programs to the household members.

#### TO OTHER RESEARCHERS

further studies are recommended for a countrywide assessment since our study only included the population of a single district (Huye). We also recommend non-cross-sectional research to determine the real origin of the infection.

## Data Availability

All data produced in the present work are contained in the manuscript

